# Nurses and Midwives’ Perspectives on Technology-Enhanced Learning and Continuous Professional Development on Emergency Obstetric and Neonatal Care in Rwanda

**DOI:** 10.1101/2023.08.28.23294717

**Authors:** Thierry Claudien Uhawenimana, Mathias Gakwerere, Anaclet Ngabonzima, Assumpta Yamuragiye, Florien Harindimana, Jean Pierre Ndayisenga

## Abstract

**Background:** One of the targets for the third sustainable development goals is to reduce worldwide maternal mortality ratio (MMR) to less than 70 deaths per 100,000 live births by 2030. To address issues affecting women and the newborns during childbirth and postnatal period, concerted efforts from governments and their stakeholders are crucial to maximize the use of technology to enhance frontline health professionals’ skills to provide the emergency obstetric and newborn care (EmONC). However, no study has garnered nurses’ and midwives’ perspectives regarding the application of technology-enhanced learning approach to provide on-job CPDs and factors that may influence the application of this training approach in the Rwandan context.

**Methods:** The study collected data from nurses and midwives from forty (40) public health facilities in remote areas nationwide. The study applied a qualitative descriptive design to explore and describe nurses’ and midwives’ perspectives on the feasibility and acceptability of technology enhanced learning approaches such as e-learning, phone-based remote training, and other online methods to provide trainings in EmONC. Two focus group discussions with EmONC mentor, two with nurses and midwives were conducted. Twelve key informant interviews were conducted. Participants were selected purposively. In total, 54 individuals were included in this study. A thematic approach was used to analyse data.

**Results:** Nurses and midwives highlighted the need to provide refresher trainings about the management of pre-eclampsia. Most of the EmONC trainings are still provided face to face and the use of technology enhanced learning approaches have not yet been embraced in delivering EmONC CPDs for nurses and midwives in remote areas. Nurses and midwives found the first developed prototype of smartphone app training of the EmONC acceptable as it met the midwives’ expectations in terms of the knowledge and skills’ gap in EmONC.

**Conclusion:** Although the newly developed application was found acceptable, further research involving practical sessions by nurses and midwives using the developed application is needed to garner views about the ease of use of the application, relevance of the EmONC uploaded content on the app, and needed improvements on the app to address their needs in EmONC.

## Introduction

Globally, women and newborns continue to be at high risk of deaths and morbidities resulting from sub-optimal care during labour, childbirth, and postpartum period [1, 2]. The World Health Organization (WHO) reports that every day, around 817 women lose their lives as a result of preventable causes related to pregnancy and childbirth [2]. Although in the last two decades significant efforts have been made to reduce maternal mortality ratio, maternal deaths still remain high in low- and middle-income countries (LMICs); particularly in the Sub-Saharan Africa (SSA) region [3]. It has been noted that SSA accounts for roughly two-thirds (196,000) of all global maternal deaths [3]. The major causes of these deaths are pregnant-related complications known as obstetric complications such severe bleeding and infections that occur after childbirth and abortion, eclampsia and pre-eclampsia, and complications from delivery; which can be prevented if women receive optimal and timely care [4, 5]. Whatever complications that expectant women experiences can affect the baby. Particularly for newborns, the first month of life is the most vulnerable period of their survival [6]. The WHO reports that 2.4 million newborns die every year; that is approximately 6700 newborn deaths every day, and most of these deaths (75%) occur during the first week of life as a result of preterm birth, birth asphyxia, neonatal infections, and other birth-related complications [6].

Therefore, it is important to invest in interventions gearing towards maternal and neonatal health improvement to address maternal and neonatal mortality and morbidities [7]. Investing in maternal health improvement provides many benefits, including improving labor supply and productive capacity in women of reproductive age, resulting in improved household income and economic well-being of families and communities [8, 9]. It also further strengthens the capacity of the health system, since many of the maternity care investments (improving human resources through capacity building, upgrading infrastructure, strengthening logistics systems for supplies and equipment, etc.) benefit other health system components as well [8].

One of the targets for the third sustainable development goals (SDGs) to reduce worldwide maternal mortality ratio (MMR) to less than 70 deaths per 100,000 live births by 2030, with no single country having a MMR higher than 140 deaths per 100,000 live birth [10, 11]. To achieve this target, it is imperative that governments and their stakeholders invest in interventions that optimize the women’s care during pregnancy, childbirth, and postnatal period. Thus, to address issues affecting women and the newborns during childbirth and postnatal period, concerted efforts from governments and their stakeholders are crucial to enhance frontline health professionals’ skills to provide the emergency obstetric and newborn care (EmONC) [7]. Building the capacity of health-care providers (HCPs) to ensure they get the necessary skills, knowledge, and competence to manage obstetric and newborn complications through ‘in-service’ training has become a common approach. Regular in-service trainings and reorientations are recommended and are in some cases mandatory, to ensure HCPs continue to be accredited by their respective professional associations [12]. In-service EmONC training programmes should build on evidence-based learning methods including technology enhanced learning methods [12]. Technology enhanced learning has a number of benefits. First, it can mitigate some service delivery gaps that occur when nurses and midwives attend face to face trainings [13, 14]. Second, those who receive face to face trainings may not find it easy to transfer the skills they earn from such trainings [13, 14]. Third, some of the people who benefit from the face-to-face trainings may leave their places of work leaving personnel gaps [13, 14]. Fourth, technology enhanced learning are efficient because they can reach many people at a reasonable cost and at a regular time and meeting the pace and convenience of nurses and midwives [13, 14]. Lastly, need to shift from traditional way of learning new skills to the new way of acquiring skills using technological tools at nurses’ and midwives’ reach; promoting efficient and effective use of resources and also bolstering their capacity in digital health [15].

Although e-learning methods have not been commonly practiced in Rwanda in the past years, the country is speeding up its pace and introducing e-learning technologies in several sectors, particularly in the health sector. In particular, e-learning and remote training is cost-effective compared to traditional learning methods and can provide space for easy scale-up and improvement of content availability [16]. Moreover, e-learning has proven its relevance in teaching and learning process amidst COVID-19 pandemic, including during challenging times of movement restrictions and lockdowns[17]. Considering the benefits of e-learning to train HCPs, there is a drive to promote technology-enhanced learning among working HCPs to build their capacity; particularly in managing some of the major issues affecting the lives of the mothers and babies during childbirth and postnatal period. For instance, UNFPA in partnership with the Ministry of Health is working to find technology based innovative ways to enhance the skills of HCPs in managing complications affecting women and their newborns during childbirth and postpartum leading to mortalities and morbidities. Under this partnership there is ongoing initiative to introduce a cost-effective and innovative solution to promote a phone-based remote training of HCPs with a particular focus on provision of EmONC services. Like other health professionals, nurses and midwives are required to continually upgrade their skills and knowledge through continuous professional development (CPD) credits. Nurses and midwives in Rwanda can obtain these CPD trainings through different avenues including face to face trainings and online trainings offered by the World Continuing Education Alliance (WCEA) and other partners. Despite the progress made, less is known about how remote nurses and midwives are facilitated to attend CPDs. It is in this regard that the current study sought to test the feasibility of providing CPDs to nurses and midwives in hard-to-reach health facilities in Rwanda using user-friendly phone-based technology. According to the successes recorded in settings where these methods were applied (in Rwanda and in other countries), the solution would ideally provide practical training through a smartphone application [18, 19]. However, in-depth user research is fundamental to provide a human-centered approach and ensure accessibility, affordability and sustainability of this technological learning approach. The United Nations Population Fund (UNFPA) Rwanda in collaboration with the Ministry of Health designed the prototype of the resolution, and it has tested it within the user research and collected user-centred feedback. Despite prospects and potential of this initiative, there has been no research conducted to assess nurses’ and midwives’ experiences with the new alternative for education.

Furthermore, available research from Rwanda has focused on health facility managers’ perceptions on the use of e-learning in providing CPD for health providers’ capacity building purposes [20]. However, to date, no study has garnered nurses’ and midwives’ perspectives regarding the application of technology-enhanced learning approach to provide on-job CPDs and factors that may influence the application of this training approach in the Rwandan context. Therefore, this study sought to understand from the nurses’ and midwives’ perspectives how technology-enhanced learning can be used to provide learner-centred CPD trainings for nurses and midwives about EmONC and factors that may influence the application of this training approach from the Rwandan context based on the newly developed EmONC smartphone application prototype.

## Methodology

### Study area

Data was collected from nurses and midwives from forty (40) public health facilities nationwide. Participating facilities included health posts, health centres, and district hospitals located in remote areas of Rwanda. Findings presented in this paper consists of the qualitative component of a countrywide mixed methods study that sought to determine how nurses’ and midwives’ access CPD for their capacity building in EmONC provision.

### Study design

This study was descriptive qualitative design [21–23]. Given that there is limited research about the use of technology enhanced learning to deliver EmONC capacity building in the Rwandan context, qualitative descriptive design enabled flexibility in data collection and analysis to garner rich comprehensive insights from nurses and midwives. This study enabled the research team to explore and describe nurses’ and midwives’ perspectives on the feasibility and acceptability of technology enhanced learning approaches such as e-learning, phone-based remote training, and other online methods to provide trainings in EmONC.

### Study population

The target population for this study was nurses and midwives working in maternity services and who were providing EmONC at the time of data collection. All nurses and midwives who worked in sampled health facilities were included in the study regardless of their experience, knowledge, and practice levels in using technology. The study excluded all participants with a work experience less than 6 months at a given health facility as they may not yet have become familiar with the facility systems and various training opportunities for EmONC.

### Sample size and sampling strategy

A random sample of one district hospital per each administrative province was selected to make 5 District Hospitals which also represented 5 administrative districts. In each district where the District Hospital was not sampled, a Health Center and Health Post were randomly selected to make 11 Health centers, and 10 Health Posts. In order to maximize qualitative feedbacks, two focus group discussions (FGDs) with EmONC mentors and two FGDs with nurses and midwives were conducted. Each FGD comprised ten members. In order to garner rich data, we also conducted five key informant interviews (KIIs) with nurses, five KIIs with midwives and, two joint interviews (one nurse and one midwife working in maternity). Participants who participated in FGDs and KIIs were selected purposively. In total, 54 individuals were included in this study.

### Data collection process

A topic guide with questions covering familiarity in using technological tools to access EmONC trainings, factors influencing technology enhanced learning among nurses and midwives, and views about the newly developed smartphone application for EmONC training was used to collect data. To garner views about the newly developed EmONC training materials featured into the app, we presented the components of the application to the participating team then we asked what they thought about them to find out once this application is launched it could be acceptable and feasible for use among nurses and midwives. Due to Covid-19 pandemic containment measures that imposed restricted movements, the research team collected data virtually as a mitigation measure for the risks that would be involved in collecting data face to face. Virtual data collection using phone calls and google meet with the support of the BEmONC mentors who are based in the data collection site was performed. EmONC district-based mentors facilitated qualitative data collectors to reach the respondents. We ensured that HCPs) felt comfortable expressing their views on the issues around the current online methods of capacity building. An experienced moderator facilitated the discussion with the assistance of a note-taker using a pre-designed interview guide. All interviews were audio-recorded. In case a respondent refused to be audio-recorded, the discussions/ interviews were written down by note-taker. Supervisors were around supervising the procedure and intervening in any case needed.

### Data analysis

The audio recorded interviews were transcribed, cleaned and translated from Kinyarwanda to English, the local language in which the discussions were conducted. To ensure the accuracy of the text, moderators and field note-takers performed the transcription. The Qualitative Research Experts performed quality checks of the transcribed and translated files, so that it accurately captures the information shared by respondents in its context. They were reading each transcript while listening its full audio and correcting errors or reviewing technical terms.

A thematic analysis was conducted and all five steps of qualitative-data analysis (reading, interpreting, coding, reducing, and displaying) were performed to ensure consistency within the data. Upon completion of transcription and synthesis of FGDs and interviews, we developed a codebook, which framed the key themes from the data. The codebook was used to systematically code a subset of FGDs and interviews transcripts. The research team (experienced moderators and note takers) and Qualitative Research experts discussed the fieldwork experience and preliminary results of fieldwork. From these discussions, a further list of codes was developed to have a final codebook that was used by qualitative research experts to analyze all qualitative data systematically. We imported the translated interviews into qualitative analysis Atlas Ti software for coding. After development of the code list, the qualitative research experts reviewed all transcripts and extracted quotes appropriate to each of the primary codes. Subsequently, the qualitative research experts reviewed all coded text and compiled themes that emerged from the data into tables with notations about where to find supporting text.

### Measures undertake to ensure trustworthiness

To ensure the rigor in the conduct of this study, interviews were conducted by experienced qualitative researcher who, in addition to their experience received a training before data collection started. Study participants were selected from different health facilities to ensure credibility through data triangulation. Furthermore, member checking was conducted to share the initial findings with participants and ensure that the researchers’ interpretation reflected the perspectives from participants and had an opportunity to clarify meaning and gather more data.

### Ethical consideration

This research was conducted after getting the ethical approval from Rwanda National Ethical Committee and the permission from Rwandan Ministry of Health (MoH) to conduct this study (Ref: NHRC/2021/PROT/036) in identified settings.

Data collectors adhered to principles of confidentiality and ethics in data collection. No person’s name (except for identification of data collector) was recorded on any of the interviews. Permission to enter each facility, to interview the different employees, and to review registers was requested from the director or staff in-charge of the health facility at the beginning of each visit. The data collection teams carried with them official letters of cooperation from the MoH and district level offices and interviewees were requested to read an information note and sign a consent form prior to proceeding with the interview. No incentive was provided to participants hence the participation was totally voluntary. Respondents were able to withdraw their participation anytime during the interview. Since the data collection was done virtually, participants first confirmed their voluntary participation by first providing a verbal consent. They were also requested to sign and return the consent form to the researchers electronically using email.

### Findings

Four major themes emerged from the findings. They encompassed i) access to EmONC CPDs, ii) experience in using technology to access EmONC information, iii) moderators of technology enhanced learning for EmONC CPD delivery, and iv) acceptability and feasibility of the newly developed technology enhanced learning ptototype for EmONC training.

### Theme 1: Access to EmONC CPDs

Most nurses and midwives reported that they found it challenging to access CPD credits online because of insufficient skills in IT and lack of continued access to the strong internet network. According to most participants, this issue affected their accumulation of required number of CPD credits in order to get their professional licenses renewed. The respondents suggested that CPD credits awarded through online system should be increased as it takes time to finish the modules then afterwards few credits are awarded as explained by one of the participants.

The respondents reflected on some of the EmONC CPD trainings they have attended but which were predominantly provided face to face. The majority of respondents attended trainings on helping mother survive and helping baby breathe (HMS and HBB respectively). The common finding among all interviewees is that they appreciated the organizers of these CPD trainings, as they included the practical sessions as reported by one of the respondents.

> *"There was a training on EmONC we had two weeks ago. We were trained on helping baby breathe and help mother survive skills. We learned how to conduct newborn resuscitation especially the positioning of newborn as someone may position the baby in a wrong way and this worsens his/her conditions. You get that it would have been more challenging to demonstrate this through online training." One midwife reported.*

Her views were echoed by most of the nurses and midwives during the FGDs.

> *"I attended training on helping mother survive. This training helped me to learn how to manage postpartum haemorrhage, severe preeclampsia, and eclampsia. This helps me a lot in my daily work. Now I am confident in what I do. I know all the steps I can take to manage those cases thanks to the practical sessions we held."*

Participants expressed ambivalent views concerning the approach that can be used to on-job EmONC CPD courses. The majority of nurses and midwives preferred face-to-face trainings over online delivered courses. They argued that physical interactions between CPD providers and learners enhances the mastery of some the topics requiring practical skills. The respondents mentioned that despite the usefulness of online CPD trainings particularly during emergencies such as Covid-19 pandemic, in-person trainings were perceived effective in equipping nurses and midwives with knowledge and translational practical skills in EmONC as illustrated in this quote.

> *"Even if we would have online CPD trainings, the face-to-face trainings are still very important. This is because face-to-face training helps learners to master the subject easily. Face-to-face learning should be maintained for some modules especially for new topics. It would be better for us to use face-to-face training because it helps people to understand courses better than with the online system."*

### Theme 2: Experience in using technology to access EmONC information

Most nurses and midwives mentioned that they have some level of experience to locate online information and resources related to EmONC. They revealed that at some point during their work, they visited websites such as Healthline and Medline to access needed information about EmONC. Participants also shared that they accessed information from their peers using the WhatsApp social media platform.

> *"In terms of access to information, especially those related to EmONC, there are many groups we have created for this purpose. For example, there is a group for all people working in maternity where we share information and this helps us. It is also used to share information about upcoming trainings on EmONC. We also discuss difficult cases on that group."*

However, some nurses and midwives; particularly from upcountry, found it challenging to search and retrieve online information about EmONC, because for some of them, the exercise required some level of knowledge in digital literacy skills and mastering all EmONC concepts in order to search relevant information meeting their needs.

### Theme 3: Moderators of technology enhanced learning for EmONC CPD delivery

Nurses and midwives reported some moderators of technology enhanced learning approaches for accumulation of EmONC CPD credits. Whilst face to face training was the most preferred, some participants were aware of the benefits they might gain when in using technology enhanced learning methods. Some of the benefits mentioned include acquiring updated skills and information on EmONC in a timely way, having access to different didactic materials including videos and slides, accessing help on some case management remotely. They also mentioned that digital based trainings are cheap and convenient for the continuity of work.

> "*What motivates me is that you get access to many things in a short time. The second thing is that you can access lessons in any format you want like Videos, audio and slides. When you cannot read slides, you use videos that demonstrate a certain procedure you want to learn. The other thing is that you can easily exchange with others about cases without having to move for face-to-face meetings*" mentioned by a respondent from KII with Midwife.

Another nurse also added that: "*It is easy to learn online as it is quick and I do it while I am at my working place. There are no transport fees required*."

The majority of respondents underscored that online learning had a potential to equip them with needed knowledge and skills to enable them to manage correctly all EmONC cases they receive thereby improving the quality of care given to mothers, babies, and families. However, participants expressed that the number of topics they covered regarding EmONC was not sufficient. They suggested that in the EmONC training package, there is a need to consider topics such as ultrasound use, management of hypertensive disorders in pregnancy, management of eclampsia, and performing manual vacuum aspiration in management of abortion. Most respondents from health centers appealed for training in cervical tear management, neonatal resuscitation in case of fetal distress, and management of Rhesus incompatibility in pregnancy it can go unnoticed quickly leading to repetitive abortions.

> *“The topic I would also like to be trained on is ultrasound. I would also like those topics about the management of preeclampsia and eclampsia be kept in the training. I am not saying that we did not receive any training on them, but when we are dealing with real cases, sometimes, it is difficult for us".* One mentor mentioned.

Another determining factor for nurses’ and midwives’ use of technology for self-directed learning about EmONC was the motivation to constantly enhancing their knowledge and skills in so that they reach a level where they felt confident and comfortable to provide quality care to mothers and newborns successfully with minimal supervision. Some of the views of the participants are shown below:

> *"For me, I want to attain a level whereby I would be able to manage any conditions that a mother and her babies might have to prevent death. This is saddening to lose a mother or a baby."* One midwife mentioned during the KIIs.

The majority of the respondents mentioned that technology enhanced learning for nurses and midwives would be successful if those providing trainings target the appropriate time for their self-learning particularly when they are off duties such as during weekends and evening hours. A few of them said that they might secure some free time to use at work when they do not have patients to attend to, especially when they have a challenging case to manage. Another alternative time for learning suggested by the respondents is when they are on annual leave.

### Theme 4: Acceptability of the newly developed ptototype e-learning app for EmONC training

Although the prototype self-learning phone application to deliver EmONC was a new concept for most of them, nurses and midwives expressed an interest in using it. They labelled it a perfect platform that will facilitate them to obtain essential skills in addressing some of the challenges they face in their daily professional activities. Most of them found it as an updated platform on current EmONC protocol. One key component most participants appreciated regarding the developed application was the ease of use and the fact that courses uploaded on it were leading to CPD credits.

> *"I think it is good, especially in terms of career development. As you said, it would be great if we could accumulate CPD credits after we have used it. This would be a good way of keeping us up to date."*

*"Personally, as a midwife, this application is needed. This is a good platform to help HCPs to stay up to date with the latest science. It will also help the HCPs to be able to manage obstetric emergencies."*

All participating nurses and midwives expressed that the digital based self-learning application came in due time because self-directed learning approach fits their nature of work. They reported that a digital based learning method would enable them to get acquainted with emerging technological advances in healthcare provision and further expose them to updated evidence-based knowledge and skills to provide maternal and neonatal care.

> *"The application is coming on time and it is a good one. What I can add on top of what have been said, is that this application will help us to continue to provide better service to our clients, as there are times when we are overworked because some of us have gone to training. We will use it while we continue to work which will help us to provide better services leading to prevention of maternal and child deaths."*

During the discussions, nurses and midwives proposed to also avail an offline option of this prototype in order to mitigate the connectivity issue that may affect its functionality. Another barrier participants reported was financial constraint to access internet bundles and smartphones for some nurses and midwives, which can reduce their interest in using the application to learn.

> *"Nurses and midwives need the offline mode because often times when it comes to internet; some people find it expensive. Others find it unnecessary to buy internet bundles for home use as they reside in areas where they have poor connectivity. For these reasons, an offline mode is better."*

> *"Before mentioning the issue of buying internet, many nurses do not even have a smartphone. Even those who have smartphones, they are not able to buy internet bundles. Therefore, an offline mode would be ideal so that they use it when they are at home and then connect themselves when they are at work using office internet."*

In terms of ways to utilize this prototype, all participants suggested there should be a way of coordinating its use at each health facility. They expressed also the need of having regular mentorship visits so that they can ask questions on what the application was not able to clarify.

*Respondent said: "Using technology and we share a link for discussing what we do not understand well in the prototype as we are doing today would be great. If possible, we could also have some time we meet face to face."*

## Discussion

We collected nurses’ and midwives’ perspectives on their capacity building needs in EmONC and how technology-enhanced learning can be maximized to bolster CPD trainings for nurses and midwives working in remote areas about EmONC. We also learnt from nurses’ and midwives’ perspectives the facilitators of and constraints that are mostly likely to influence the application of technology enhanced learning approach in training nurses and midwives working in remote areas about EmONC.

We found that access of CPDs by nurses and midwives still need to be improved because they still do not get all needed EmONC skills based on their personal work experiences. Particularly, our study found that there is a need to provide refresher trainings about the management of pre-eclampsia. The findings from this study corroborate those of studies conducted from different low resources countries across the globe [24–27], that established the need for providing adequate and regular trainings on EmONC for nurses at primary health level to enable them recognize the risk factors of preeclampsia, and be able to confirm it so that prophylactic interventions to manage it are initiated earlier. This finding also supports the results of a similar study conducted in Rwanda that highlighted lack of regular and continuous trainings for nurses and midwives about EmONC as a constraint to managing emergency maternal and neonatal complications including pre-eclampsia [27].

The current study established that most of the EmONC trainings are still provided face to face and the use of technology enhanced learning approaches have not yet been embraced in delivering EmONC CPDs for nurses and midwives in remote areas. There are several explanations for this result. First, this may be due to individual and systemic factors including the limited familiarity of health providers familiarity with the internet and computer usage as well as low familiarity with online methods of teaching learning. Preference to be trained face to face with the trainer and financial benefits that may be associated with attending face to face trainings. Second, systemic factors: limited IT resources at the facilities and their readiness to adopt online learning approaches in building the capacity of their staff. This is confirmed by a study that involved managers of health facilities that identified challenges such as the lack of access to digital devices, poor or lack of internet access, poor online learning design, low digital skills of healthcare professionals, lack of time dedicated to online learning, and heavy workload of staff as barriers affecting the provision of CPD and capacity building trainings for health professionals using technology enhanced learning approach [20].

Nurses and midwives are willing to undertaken EmONC CPDs delivered through blended approach using both online approach and hands-on-skills physical trainings. Our findings support those of a study conducted by Nyiringango et al. in which nurses, midwives, and physicians perceived the need for blended mode of CPD provision [28].

Nurses and midwives found the first developed prototype of smartphone app training of the EmONC acceptable as it met the midwives’ expectations in terms of the knowledge and skills’ gap in EmONC. This finding is consistent with perspectives expressed by midwives in Tanzania about the effects of smartphone application in equipping them with knowledge useful for their practice of EmONC provision [29]. In spite of the positive perspectives about the acceptability of the newly developed smartphone app, our study, identified a number of constraints that may affect nurses’ and midwives’ utilization of the newly developed smartphone application. Nurses and midwives mentioned that the quality of and availability of internet in the remote areas, affordability of internet bundles, and the coordination of the online training for CPD purposes may affect the use of this application for CPD purposes. Like in other places where smartphone applications were piloted to provide trainings such as Tanzania, similar constraints above mentioned have been documented particularly lack of free internet access in maternity [29]. In addition to these challenges, unlike our study, the study from Tanzania further highlighted lack of time to read due to busy workloads [29]. To mitigate the issue of limited internet network access, nurses and midwives suggested that they would actively use the app to access CPDs on EmONC if an offline version of the app is availed. Our study further suggested that regular monitoring to check the utilization of the app will need to be carried to support nurses and midwives who may experience difficulties whilst using the app.

## Limitations

The current study has some limitations that may call for cautious interpretation of the findings. Participants were asked their views about the feasibility and acceptability of the application they have seen through a brief demonstration but which they have not used before. For this reason, we could not garner participants’ views on the ease and/difficulty of using the application in accessing EmONC CPD materials. Could they have got enough time to test and use the newly developed application and accessing the content of the application, the study could have garnered more insights on the feasibility of the developed application. Another limitation is that data was collected virtually due Covid-19 pandemic containment measures. Although we tried our best to minimise any biases, the involvement of district based EmONC mentors to facilitate the smooth running of interviews may have led to some respondent biases. Lastly, although the current study sought to explore how online and computer-based CPD delivery can be promoted to provide CPDs to nurses and midwives, it cannot be ignored that nurses and midwives need hand on skills practice to provide EmONC to the mothers and newborns hence a blended approach would always add value as highlighted by some respondents in this study.

## Conclusion

Based on the results from this study, the use of technology enhanced approach to provide EmONC CPDs to nurses should be encouraged. From the nurses’ and midwives’ perspectives, technical themes that need more attention in regards to CPDs on EmONC for nurses and midwives from remote health facilities in Rwanda have been highlighted. Based on the collected views, there is a need to equip nurses and midwives with skills about detection and management of pre-eclampsia and further provide advanced capacity building trainings for nurses and midwives in other areas of EmONC.

The study found that nurses and midwives have not yet maximized the use of internet for self-learning purposes. In addition, the use of online resources such as academic websites by nurses and midwives for self-learning is still very limited. Therefore, interventions to train nurses and midwives in digital health literacy such as searching for information, appraising it, and selecting what content is relevant to enhancing their skills in EmONC are needed.

The study established that nurses and midwives may undertake capacity building trainings online if the content provided respond to their gaps in knowledge and practice challenges encountered regarding the provision of EmONC. In addition, to meet the nurses’ and midwives’ expectations, the content provided online and/or using technology enhanced learning approaches need to take into consideration their workload and availability.

Although the newly developed application was found acceptable, further research involving practical sessions by nurses and midwives using the developed application is needed to garner views about the ease of use of the application, relevance of the EmONC uploaded content on the app, and needed improvements on the app to address their needs in EmONC.

## Data Availability

Data pertaining to this study have been included in the manuscript. Additional records and transcripts can be obtained from the research team upon writing to the submitting author.

## References

1. Kassebaum NJ, Barber RM, Bhutta ZA, et al (2016) Global, regional, and national levels of maternal mortality, 1990–2015: a systematic analysis for the Global Burden of Disease Study 2015. Lancet 388:1775–1812

2. Organization WH (2019) Trends in maternal mortality 2000 to 2017: estimates by WHO, UNICEF, UNFPA, World Bank Group and the United Nations Population Division.

3. WHO (2023) Trends in maternal mortality 2000 to 2020: estimates by WHO, UNICEF, UNFPA, World Bank Group and UNDESA/Population Division. Geneva

4. Say L, Chou D, Gemmill A, Tunçalp Ö, Moller A-B, Daniels J, Gülmezoglu AM, Temmerman M, Alkema L (2014) Global causes of maternal death: a WHO systematic analysis. Lancet Glob Heal 2:e323–e333

5. Musarandega R, Nyakura M, Machekano R, Pattinson R, Munjanja SP (2021) Causes of maternal mortality in Sub-Saharan Africa: a systematic review of studies published from 2015 to 2020. J. Glob. Health 11:

6. WHO (2023) Newborn mortality: Key facts. https://www.who.int/news-room/fact-sheets/detail/levels-and-trends-in-child-mortality-report-2021#:~:text=Preterm birth%2C intrapartum-related complications,causes of most neonatal deaths.

7. Organization WH (2021) Ending preventable maternal mortality (EPMM): A renewed focus for improving maternal and newborn health and well-being.

8. Sully EA, Biddlecom A, Darroch JE, Riley T, Ashford LS, Lince-Deroche N, Firestein L, Murro R (2020) Adding it up: investing in sexual and reproductive health 2019.

9. Remme M, Vassall A, Fernando G, Bloom DE (2020) Investing in the health of girls and women: a best buy for sustainable development. BMJ 369:m1175

10. World Health Organization (2018) Maternal mortality fact sheet.

11. Howden-Chapman P, Siri J, Chisholm E, Chapman R, Doll CNH, Capon A (2017) SDG 3: Ensure healthy lives and promote wellbeing for all at all ages. A Guid to SDG Interact from Sci to implementation Paris, Fr Int Counc Sci 81–126

12. Ameh CA, van den Broek N 2015) Making It Happen: Training health-care providers in emergency obstetric and newborn care. Best Pract Res Clin Obstet Gynaecol 29:1077–1091

13. O’Connor S, Kennedy S, Wang Y, Ali A, Cooke S, Booth RG (2022) Theories informing technology enhanced learning in nursing and midwifery education: A systematic review and typological classification. Nurse Educ Today 118:105518

14. Gause G, Mokgaola IO, Rakhudu MA (2022) Technology usage for teaching and learning in nursing education: An integrative review. Curationis 45:2261

15. Ngenzi JL, Scott RE, Mars M (2021) Information and communication technology to enhance continuing professional development (CPD) and continuing medical education (CME) for Rwanda: a scoping review of reviews. BMC Med Educ 21:245

16. Wall L, Nartker A, McGee A, Scott E, Downer A (2017) No internet? No problem! Creative approaches to cost-effective e-learning delivery in resource-constrained settings. Lancet Glob Heal 5:S8

17. Jeffries PR, Bushardt RL, DuBose-Morris R, Hood C, Kardong-Edgren S, Pintz C, Posey L, Sikka N (2022) The role of technology in health professions education during the COVID-19 pandemic. Acad Med 97:S104

18. Emmanuel G, Emanuel A, Setyohadi D (2020) Design of mobile application for community health workers: A case study in Rwanda.

19. Rusatira JC, Tomaszewski B, Dusabejambo V, et al (2016) Enabling Access to Medical and Health Education in Rwanda Using Mobile Technology: Needs Assessment for the Development of Mobile Medical Educator Apps. JMIR Med Educ 2:e7

20. Byungura JC, Nyiringango G, Fors U, Forsberg E, Tumusiime DK (2022) Online learning for continuous professional development of healthcare workers: an exploratory study on perceptions of healthcare managers in Rwanda. BMC Med Educ 22:851

21. Denzin NK, Lincoln YS (2003) The landscape of qualitative research : theories and issues. J Adv Nurs 51:xi, 684 p.

22. Nseir S, Le Gouge A, Pouly O, et al (2021) Relationship Between Obesity and Ventilator-Associated Pneumonia: A Post Hoc Analysis of the NUTRIREA2 Trial. Chest 159:2309–2317

23. Doyle L, McCabe C, Keogh B, Brady A, McCann M (2019) An overview of the qualitative descriptive design within nursing research. J Res Nurs 25:443–455

24. Atluri N, Beyuo TK, Oppong SA, Moyer CA, Lawrence ER (2023) Challenges to diagnosing and managing preeclampsia in a low-resource setting: A qualitative study of obstetric provider perspectives from Ghana. PLOS Glob Public Heal 3:e0001790

25. Seif SA, Rashid SA (2022) Knowledge and skills of pre-eclampsia management among healthcare providers working in antenatal clinics in Zanzibar. BMC Health Serv Res 22:1512

26. Garti I, Gray M, Tan J-Y, Bromley A (2021) Midwives’ knowledge of pre-eclampsia management: A scoping review. Women and Birth 34:87–104

27. Nishimwe A, Conco DN, Nyssen M, Ibisomi L (2022) Context specific realities and experiences of nurses and midwives in basic emergency obstetric and newborn care services in two district hospitals in Rwanda: a qualitative study. BMC Nurs 21:9

28. Nyiringango G, Byungura JC, Fors U, Forsberg E, Tumusiime D (2023) Online learning needs, facilitators, and barriers for continuous professional development among nurses, midwives, and physicians in Rwanda. Int J Africa Nurs Sci 18:100574

29. Shimpuku Y, Mwilike B, Mwakawanga D, Ito K, Hirose N, Kubota K (2023) Development and pilot test of a smartphone app for midwifery care in Tanzania: A comparative cross-sectional study. PLoS One 18:e0283808

